# Novel method for estimating right atrial pressure with point-of-care ultrasound (POCUS)

**DOI:** 10.1101/2022.05.05.22274742

**Authors:** Larry Istrail, Joseph Kiernan, Maria Stepanova

## Abstract

Current noninvasive estimation of right atrial pressure (RAP) by either bedside jugular venous pressure (JVP) exam or inferior vena cava (IVC) measurement during a formal echocardiogram offer imprecise estimates of actual RAP. We enrolled 41 patients in a prospective, blinded study to validate a novel point-of-care ultrasound method to estimate RAP. Two subjects were excluded and 39 were included in the final analysis. The ultrasound estimate of RAP (RAP_U_) was compared to the RAP measurement during right heart catheterization (RAP_i_) both as measured and corrected for the mid-AP diameter. The correlation coefficient between RAP_i_ and corrected RAP_U_ measurements was +0.72, regression R^2^ 0.52, bias −0.60 mmHg (95% confidence interval [CI], −1.60 to +0.39 mmHg) with the limits of agreement −5.56 to +7.24 mmHg, and 3 mmHg accuracy of 26 (67%). Similarly, for the uncorrected RAP_U_ measurement, the correlation coefficient was +0.75, regression R^2^ 0.56, bias −0.49 mmHg (95% CI, −1.42 to +0.43 mmHg) with the limits of agreement −5.56 to +7.24 mmHg, and 3 mmHg accuracy of 29 (74%). This simple bedside evaluation of right atrial depth and the right jugular vein correlates with actual right atrial pressure better than traditional IVC parameters, and can accurately estimate RAP within 3mmHg in most patients.

## Background

Noninvasive estimation of right atrial pressure (RAP) is currently done by either bedside jugular venous pressure (JVP) exam or by measuring inferior vena cava (IVC) diameter and collapsibility during a formal echocardiogram. However, both of these measures offer imprecise estimates of actual RAP. The visual inspection of the jugular vein first described by Sir Thomas Lewis in 1930^1^ has been the basis of our volume exam. However, it has many limitations that result in poor sensitivity and low diagnostic accuracy, such as the inability to visualize the jugular vein in many patients with thick necks or who are obese.^2–8^ This method also assumes the right atrium is 5 centimeters below the sternum based on data from a 1946 study of chest x-rays,^9^ when in fact it can vary from 5cm to 15 cm.^10,11^

Echocardiographic IVC measurement to estimate RAP is the current non-invasive gold standard, however, it only offers a very weak correlation to actual RAP. In a blinded, prospective study, Magino et al found that both IVC diameter and collapsibility had R^2^ of 0.19 or less, and was within 2.5 mmHg of the actual value only 34% of the time.^12^ These inaccuracies lead to overestimations in pulmonary pressures, can misguide diuretic treatment choices, and ultimately lead to clinical uncertainty and invasive right heart catheterizations to make the final determination of volume status.^13^

Point-of-care ultrasound (POCUS) offers alternative methods for estimating RAP at the bedside. It can overcome the anatomical limitations present in visually inspecting the jugular vein and can also measure the right atrial depth (RAD) in each patient. Taken together, these two measurements may provide a more accurate, non-invasive, and quantitative measurement of right atrial pressure.

## Methods

A prospective, blinded study was completed to compare the performance of a novel internal jugular vein ultrasound technique to estimate right atrial pressure. Patients undergoing RHCs were recruited for this study if they were 21 years old or over and required RHC for any purpose. Patients were excluded from the study based on the following: 1) right internal jugular vein catheter present; 2) intubated or on positive pressure ventilation; 3) congenital heart disease history; 4) unable to visualize the posterior LVOT in parasternal long-axis view. All patients provided written informed consent before enrollment in the study. The study was approved by the IRB.

Within 2 hours prior to the right heart catheterization (RHC), the patient’s head of the bed was placed at 45 degrees and one physician certified in POCUS (LI) performed the ultrasound exam. A Butterfly IQ+ probe was placed along the left sternal border and a cardiac parasternal long-axis view was acquired. With the probe positioned as close to perpendicular to the chest wall as possible, the distance from the chest wall down to where the non-coronary cusp of the aortic valve attaches to the posterior wall of the left ventricular outflow tract (LVOT) was measured (Figure 1). This landmark was chosen as a surrogate for the center of the right atrium because it sits in the same approximate coronal plane (as seen in the parasternal short-axis view at the aortic valve level), and is easily detectable in a supine patient. This distance was recorded as the RAD.

**Figure 1:**
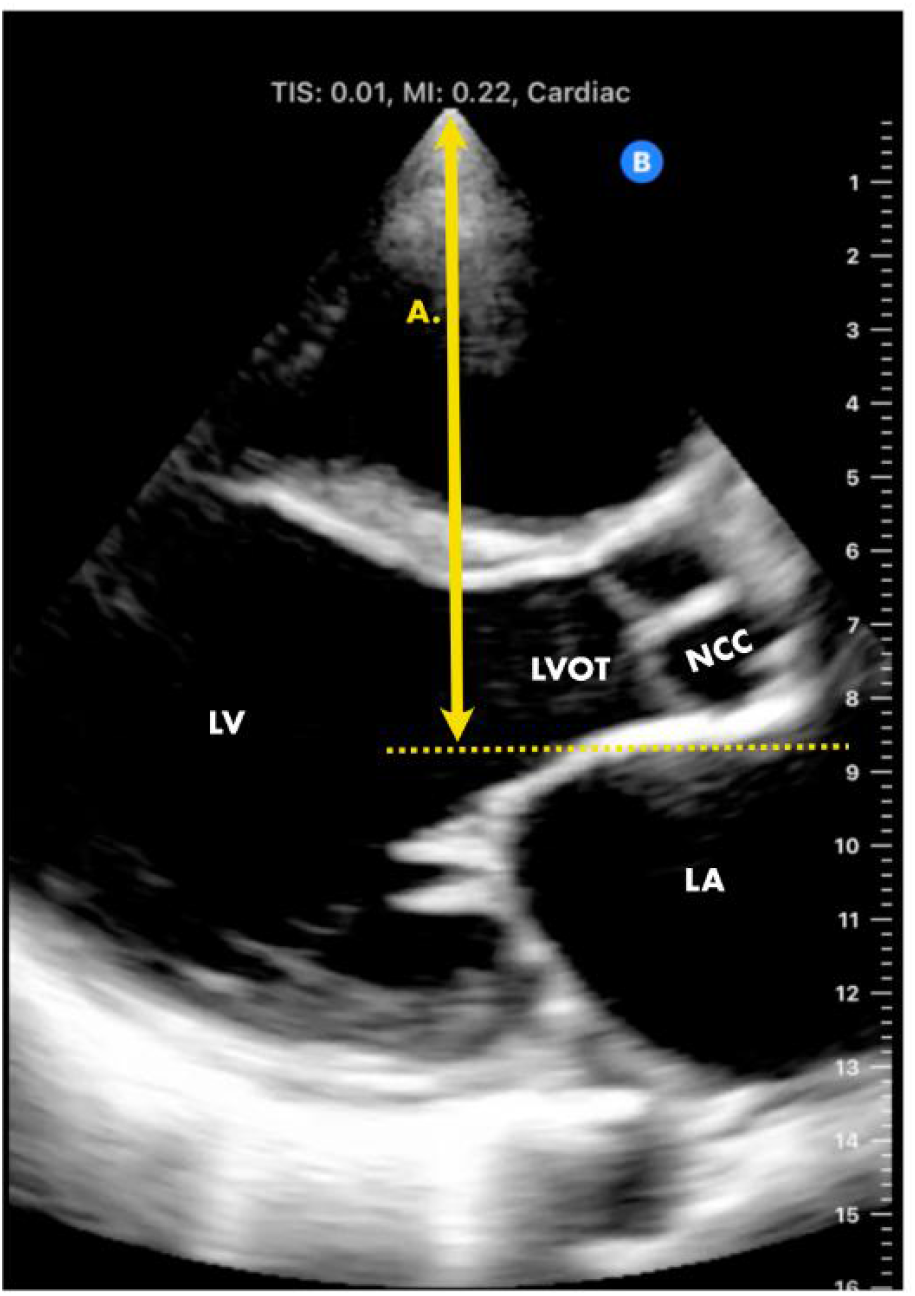
Right Atrial Depth (RAD) A. Right atrial depth (RAD). With the probe perpendicular to the chest wall, the distance from the chest wall to the posterior wall of the LVOT where the non-coronary cusp (NCC) attaches is measured.

The probe was then gently placed just superior to the clavicle at what we called the supraclavicular point. In the transverse view, the IJV was visualized, and a clip at this point was recorded. IJV shape, degree of collapsibility, or presence of jugular venous distention (JVD) was determined. JVD was considered present at the supraclavicular point if the vein was distended with minimal or absent jugular venous pulsations. JVD was deemed not present if the vein collapsed completely with normal respiration, even if the jugular vein was enlarged during inspiration.

The ultrasound estimated RAP (RAP_U_) value was calculated in two discrete ways depending on if there was JVD present at the supraclavicular point or not. If JVD was present at the supraclavicular point, the ultrasound probe was slid cranially to identify the jugular venous collapse point where the venous walls collapse completely. The probe was then rotated 90 degrees into the longitudinal plane to confirm this point. This is the meniscus of the blood column that corresponds to the jugular venous pulse (Figure 2). We will call it the ‘Wine Bottle Sign,’ due to the previous description of its resemblance to the top of a wine bottle in the longitudinal plane.^14^ The vertical distance from this point down to the sternum was then measured in centimeters using a ruler. This value was then added to the RAD in cm to estimate RAP_U_.

**Figure 2:**
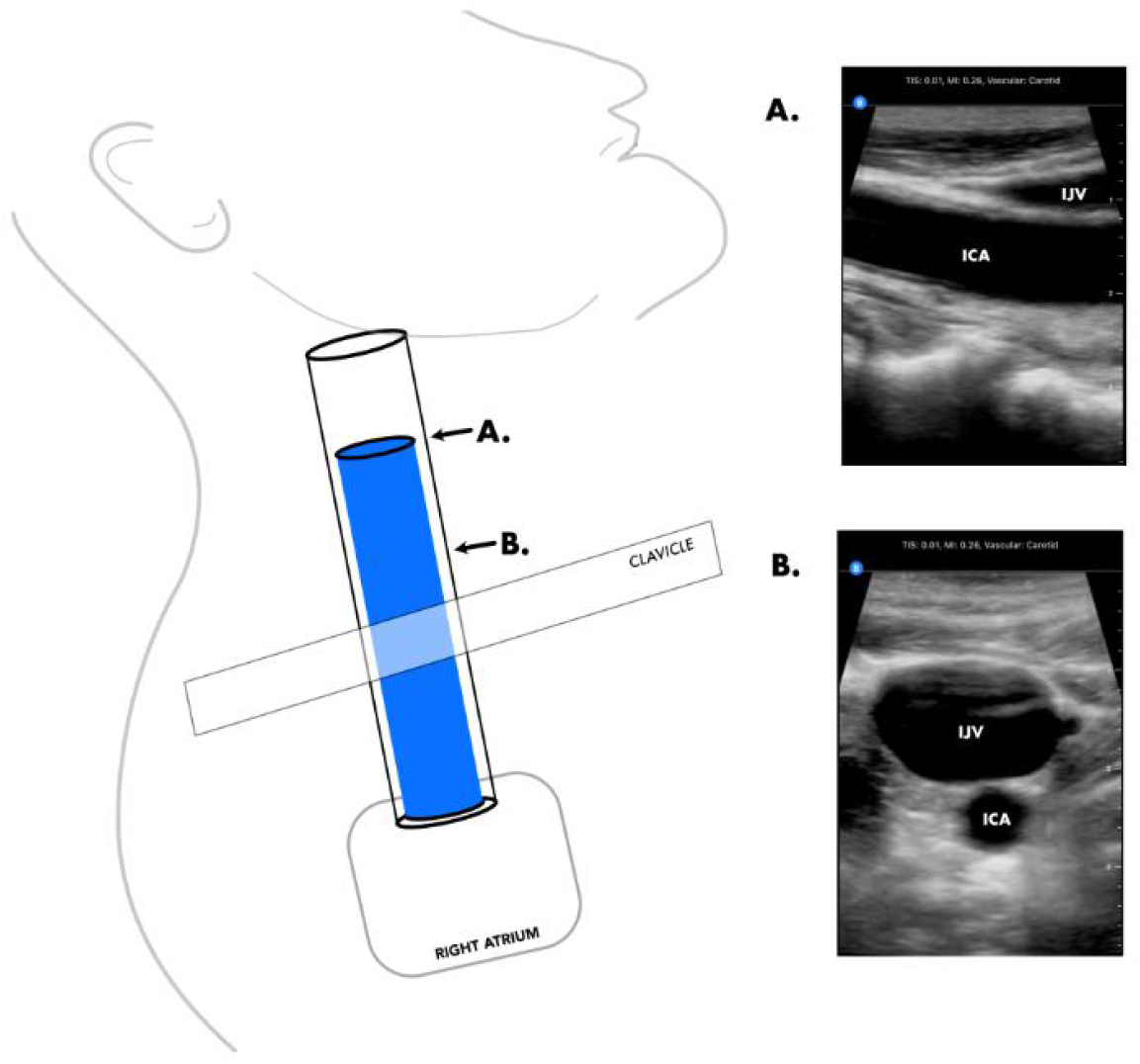
Appearance of Jugular Venous Distention and Wine Bottle Sign with POCUS. **A:** Wine Bottle Sign: Meniscus of blood column in internal jugular vein (IJV) where jugular venous pulsations most prominent. IJV assumes a triangular appearance at the meniscus. IJV lies just superficial to internal carotid artery (ICA) **B:** Presence of JVD at supraclavicular point. IJV is dilated with minimal or absent pulsations

If the IJV was completely collapsing with each breath at the supraclavicular point with the head of the bed at 45 degrees, then JVD was deemed absent. In order to estimate the pressure in this case, the angle of the head of the bed was lowered to 30 degrees and the IJV was assessed for partial or full engorgement greater than that seen at 45 degrees. If the vein did not engorge to a greater degree than at the 45-degree position, then the head of the bed was dropped further to zero degrees, and the presence of partial or full engorgement was recorded.

If the vein engorged at 30 degrees, then the RAP_U_ in cmH2O was estimated to be RAD x 0.75. If the IJV engorged at zero degrees, then the RAP was estimated to be RAD x 0.5. If the vein still did not engorge at zero degrees, then the RAP was estimated to be RAD x 0.25. The pressure in cmH2O was then converted to mmHg by multiplying by 0.735. Lastly, the anterior-posterior (AP) diameter of the chest was measured and the mid-AP diameter was marked. The estimated RAP_U_ was then recorded, as was the RAP_U_ corrected for the mid-AP diameter. Both values were then entered into the REDCap database prior to the RHC.

The RHC was then performed by one interventional cardiologist (JK) through right brachial or jugular venous access. The zero reference level was set to the measured mid-thoracic level and the invasive right atrial pressure (RAP_i_) was measured. The interventionist was blinded to the results of the ultrasound results.

### Statistical analysis

Data were presented as mean ± standard deviation or as median and interquartile range when appropriate. The associations of RAP_U_ and RAP_i_ measurements were tested using Pearson correlation and linear regression. The agreement between the two methods was evaluated by the Bland-Altman analysis, and the 3 mmHg accuracy was calculated for corrected and uncorrected ultrasound measurements separately.

## Results

Forty-one subjects were consented and enrolled in the study. Two were excluded (one was scheduled for both right and left heart cath but only underwent left heart cath, and in one the LVOT could not be visualized in the parasternal long-axis view) and 39 were included in the analysis. The mean age was 63 (SD 14) years, 49% male, BMI 28.4 (5.4) (**Table 1**).

**Table 1.** Patient characteristics (n = 39).

| Parameter |  |
| --- | --- |
| Age, years (mean ± SD) | 63.3 ± 14.1 |
| Male, % | 19 (49%) |
| BMI (mean ± SD) | 28.4 ± 5.4 |
| Invasive measurement, mmHg (mean ± SD, median (IQR)) | 5.6 ± 4.3, 5.0 (2.0 - 7.5) |
| Ultrasound measurement corrected (mean ± SD, median (IQR)) | 6.2 ± 3.9, 4.4 (3.7 - 8.8) |
| Ultrasound measurement not corrected (mean ± SD, median (IQR)) | 6.1 ± 3.6, 4.4 (3.3 - 9.0) |

The association between RAP_i_ and RAP_U_ measurements was strong whether the right atrial depth was corrected for the mid-thoracic diameter or not (**Figure 3**). Specifically, the correlation coefficient between RAP_i_ and corrected RAP_U_ measurements was +0.72, regression R^2^ 0.52, bias −0.60 mmHg (95% confidence interval [CI], −1.60 to +0.39 mmHg) with the limits of agreement −5.56 to +7.24 mmHg, and 3 mmHg accuracy of 26 (67%). Similarly, for the uncorrected RAP_U_ measurement, the correlation coefficient was +0.75, regression R^2^ 0.56, bias −0.49 mmHg (95% CI, −1.42 to +0.43 mmHg) with the limits of agreement −5.56 to +7.24 mmHg, and 3 mmHg accuracy of 29 (74%).

**Figure 3:**
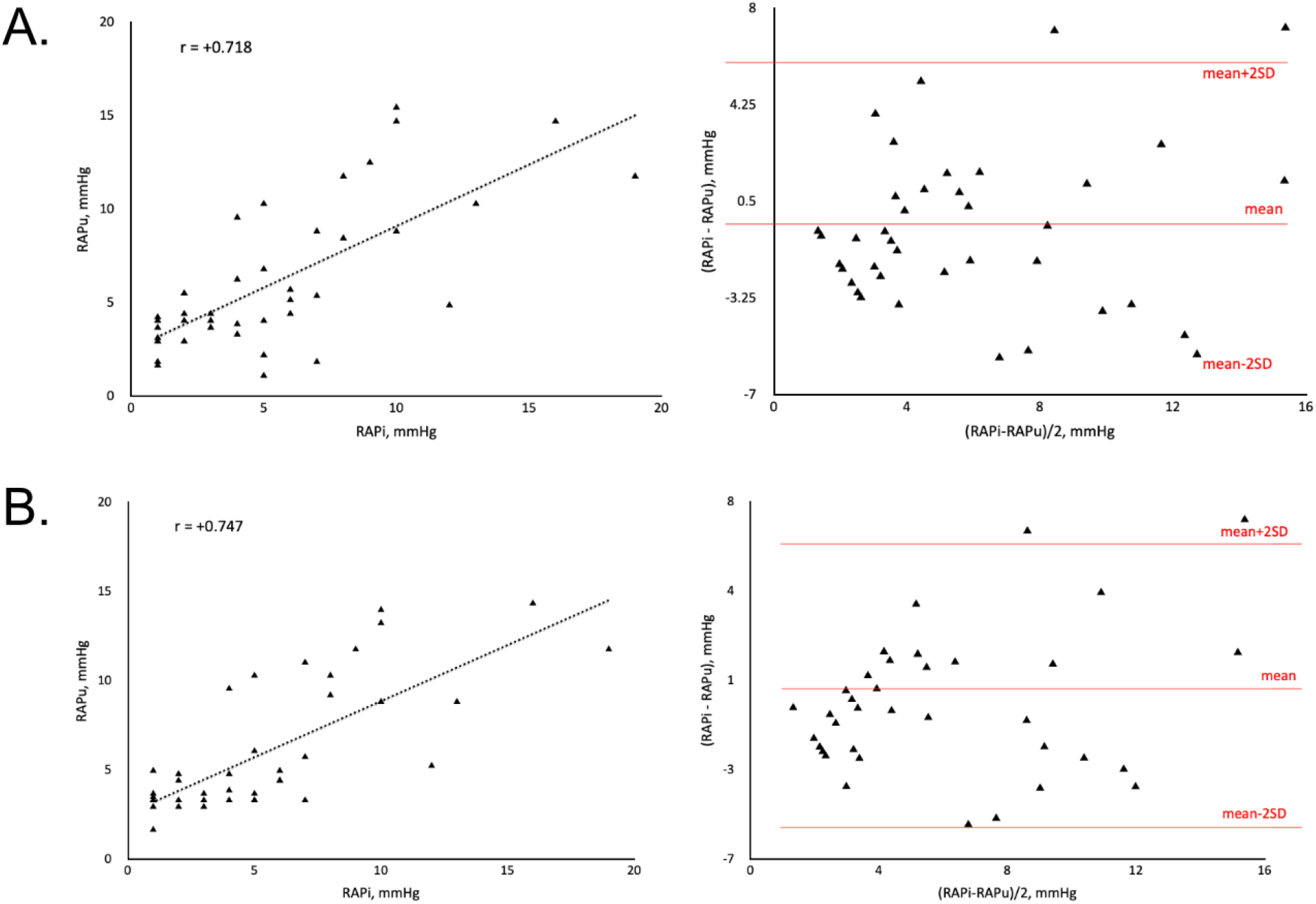
Comparison of invasive (RAPi) and ultrasound (RAPu) RAP measurements. **A:** Corrected RAPu; **B:** Uncorrected RAPu. Left: scatter plot with a linear regression line; right: Bland-Altman plot.

## Discussion

The main finding of this study is that a measurement of RAD coupled with jugular venous ultrasound exam can accurately estimate RAP regardless of patient body habitus or neck anatomy. Previously we compared this POCUS method to RAP obtained from RHC performed by multiple interventional cardiologists without standardized zeroing technique between them, resulting in less accurate results.^15^ When the RHC zero level was standardized to the mid-thoracic level, both the corrected and uncorrected RAP_U_ measurements strongly correlated with gold-standard RAP_i_ measurements, and was within 3 mmHg 74% of the time. This is compared to an R^2^ of 0.19 and overall accuracy of 34% seen with standard IVC measurements.^12^ In cases where the RAP_U_ was not accurate in predicting RAP_i_ within 3mmHg, it was always within 5 mmHg of RAP_i_ with one exception. In this exceptional case, the wine bottle sign was not visualized because the JVD extended above the mandible and thus the predicted value was entered as ‘greater than 11.7 mmHg,’ corresponding to an actual RAP_i_ of 19 mmHg.

Other studies have already shown that visualizing the jugular vein with ultrasound is reliable and feasible in every patient; however, these studies still rely on the 5-centimeter assumption for right atrial depth which may explain their underestimation of actual RAP.^16,17^ This is likely due to the fact that the RAD is on average much deeper than 5 centimeters. In our study, using the non-coronary cusp attachment to the posterior LVOT as a proxy, the average distance to the right atrium at the 45-degree position was 9.2 cm. This is similar to the RAD in our previous study of 10.16 cm,^15^ as well as to the 9 centimeters in Kovacs et al^10^ and the 9.9 cm found by Seth et al^11^ which both relied on measurements from CT scans.

Our measurement accuracy within 3mmHg was strongest with the uncorrected RAP_u_ at 74%. This suggests that when the RHC is zeroed at the mid AP diameter, correcting for the RAD may not be necessary for clinically meaningful results. This method could offer a simple and reliable RAP estimate within 3 mmHg, which would have a profound impact on bedside medicine, diuretic use, and need for RHC in patients with unclear volume status. Since IVC diameter and collapsibility was only 25-37% accurate in other studies,^12^ this method could also offer better quantitative estimates of RAP during formal echocardiograms which would improve non-invasive pulmonary pressure estimates as well.

This study was subject to several limitations. It was a single-center study using a convenience sample of patients in need of right heart catheterization for any reason. It consisted of both outpatients and inpatients without congenital heart disease but excluded critically ill patients requiring intubation or non-invasive ventilation which may have biased the results.

## Data Availability

All data produced in the present study are available upon reasonable request to the authors

